# How decentralized facility financing improved supply chains and product availability in primary healthcare centers, a randomized trial in Nigeria

**DOI:** 10.1101/2025.07.21.25331940

**Authors:** Brittany Hagedorn, Jeremy Cooper, Oluwole Odutolu, Ojukwu Mark Ojukwu, Benjamin Loevinsohn

**Affiliations:** Institute for Disease Modeling, Gates Foundation, Seattle WA USA; Poliwonk Nigeria Limited, London UK; World Bank, Abuja FCT Nigeria; Gavi, The Vaccine Alliance, Geneva Switzerland

## Abstract

**Background:** The availability of essential medicines and supplies remains a serious impediment to effective primary health care (PHC) in many lower and lower middle-income countries. Most of these countries rely on centralized procurement, centralized stores, and a “push” distribution system. We describe here the impacts of a large-scale randomized trial in Nigeria, which provided modest funding directly to facilities to spend, on supply availability.

**Methods:** Districts in three states were randomly allocated to either direct facility financing (DFF) or performance-based financing (PBF) and matched to a control group. Both DFF and PBF transferred funds to facility bank accounts and allowed the facility management committee to spend on operational costs, including essential drugs. Facilities could procure medicines on the government’s essential drug list from pre-approved suppliers if they were certified by the national drug regulator. We conducted a difference-in-difference (DiD) analysis using facility survey data to assess the impact on availability of essential drugs and supplies.

**Results:** Drug availability was initially similar for the three arms of the trial (9 of 29 essential medicines). After three years, DFF and PBF facilities had significantly higher product availability than control (p<0.05). This amounted to an increase of 28/ 34 percentage points in DFF/PBF facilities (an additional 8/10 products, respectively), and only 10% in control (3 additional products). We did note that there was little difference between control and intervention arms in the availability of medicines for donor-supported vertical programs like immunization, family planning, and malaria. However, there were very large improvements for products like antibiotics, obstetrical drugs, diagnostics, and TB medications.

**Conclusion:** Providing funds directly to health facilities improved drug availability. It was superior to the typical centralized procurement and “push” distribution system that is widespread in lower-income settings. This approach is already spreading and should be adopted more widely.

## Introduction

A reliable supply chain that can provide safe drugs and consumables to health facilities is an essential ingredient of an effective primary healthcare (PHC) system. (1) However, building out a system that can routinely deliver and ensure availability of essential products has proven elusive in many countries, especially those with limited resources such as Nigeria. (2,3) Most low and lower middle-income countries use a centralized procurement and distribution system that still fails to prevent stock-outs, despite decades of effort and significant financial investments. These supply chains typically rely on a public sector, “push” based system, which provides little agency to front-line health workers and their managers at health facility level. We describe here an alternative approach where funds were provided directly to health facilities and they were responsible for purchasing supplies and pharmaceuticals from accredited private sector wholesale pharmacies. This approach was tested on a large scale and subjected to a randomized trial.

Studies across lower and lower middle-income countries reveal that the availability of essential medicines and supplies remains a serious concern. One review across 10 countries found that only 19% of basic PHC facilities had relevant diagnostics. (4) Another study looking at the availability of essential medicines for asthma and chronic obstructive pulmonary disease found that only six of 58 (10.3%) countries studied met the WHO standard of 80% availability. (5)

Like other lower middle-income countries, Nigeria’s PHC supply chain management system faces numerous challenges, including difficulties with product distribution and inventory management (6). Essential drug availability at primary healthcare centers was below 50% nationally in a 2019 (7,8) study, a level that can compromise quality of care (9–11). When faced with limited supply, concerns about counterfeit and ineffective products can be a concern, although this risk is variable (12,13) and studies of have found a low prevalence. (14,15)

Several factors contribute to stock-outs, including poor storage infrastructure, financial constraints, insecurity, transportation challenges, inadequate human resources, and weak or poorly implemented policies as contributing causes of stock-outs in public sector facilities. (16,17) This is despite continued investment in the public sector supply chain and many pilot studies purporting to identify the right redesign approach. (18,19)

Part of the challenge is that the public sector supply chain system in Nigeria is complex, with players at the national and state levels (20), vertical disease-specific efforts and donor-funded programs. (21) The Nigerian constitution stipulates (22) that states are responsible for managing their respective primary and secondary health systems and consequently, management systems can vary widely from state to state (23), in part reflecting varying levels of health system and market development. These can include state-owned procurement and distribution systems, the use of drug management agencies, annual tender processes with accredited suppliers, and adoption of an open tender system. (16,20) Regardless of the approach, the supply chain for public facilities has historically been dominated by a government-run system that relies on central procurement and government-managed stores, and distribution barriers stemming from this are the most-cited reason for stock-outs. (3) We note that this centralized approach is not used in higher income countries that rely instead on private sector distribution.

This underscores the potential impact of testing new approaches, and many potential interventions have been examined (24–26). A variety of studies, including randomized controlled trials (RCTs) and simpler pre/post evaluations, have been used to evaluate the effectiveness of strategies aimed at improving supply chain performance and reducing stock-outs. (24–26) For example, a study in Uganda that leveraged supervision, performance assessment, and recognition improved supply chain management practices by 16 percentage points. (27) The introduction of logistics management systems in Tanzania reduced stock-outs rates from 35% to 22%, although a similar system in Ethiopia was found to be limited by human resources skills and poor implementation. (28,29) In Zambia, an RCT that improved logistics capacity at the district level and reduced the number of stockholding points showed meaningful reductions in stock-outs, with a 7-52% percentage point improvement across the six tracer drugs when the role of the district was shifted to cross-docking rather than repackaging and facilities submitted ‘pull’ requests for restocking. (30,31) These trials demonstrate the potential for improving supply chain management and reducing stock-outs, showing that significant gains can be made when decision making is shifted closer to the facility.

One approach to study possible solutions is the Nigeria State Health Investment Project (NSHIP), a Nigerian government initiative financed by World Bank credit. It aimed to strengthen the PHC system, increase the delivery of high-impact maternal and child health interventions, and improve quality of care; it did not set out to strengthen the supply chain per se. (32) Local Government Areas (LGAs equivalent to districts) in three states were randomly assigned to either direct facility financing (DFF) or performance-based financing (PBF), with all the public PHC facilities receiving incremental funding paid directly into the facility’s bank account. The facilities in the PBF arm were paid based on the number and quality of services they provided (annual average $3.49 per capita). The facilities in the DFF arm received an amount calculated as 50% of what PBF facilities were earning, unconditional on performance (annual average $1.74 per capita). In addition to the incremental funding, the PBF and DFF facilities received training on management and budgeting, enhanced supervision using a quantitative supervisory checklist, and involvement of a community representative on the health facility management committee. (35) As a result, DFF improved Penta3 vaccination coverage by 20 percentage points, decreased out-of-pocket expenditures for child curative care, and improved structural quality of care. (32)

The health facility management committees in both PBF and DFF facilities had considerable autonomy in how they used the funds they received. They could use them for e.g., repairs and maintenance, conducting outreach, or to purchase supplies and medicines. The latter had to be: (i) on the government’s essential drug list; (ii) approved by the National Agency for Food and Drug Administration and Control (NAFDAC); (iii) purchased from a manufacturer or distributor licensed by the Pharmaceutical Council of Nigeria (PCN); and (iv) from a state-approved wholesaler. Health facilities could make purchases from both government and private-sector distributors. Health authorities ensured that facilities followed the guidelines through routine supervision, including use of a checklist that tracked practices related to proper stock management.

This was in sharp contrast to the typical reliance on centralized control with Central Medical Stores and distribution with a ‘push’ system and in-kind donation of drugs through vertical programs. Instead, the DFF/PBF strategy allowed facilities to spend small amounts of money to purchase supplies as needed, creating a ‘pull’ system that allowed for planning at the facility level based on local pharmaceutical and medical consumable needs. This was a novel procurement mechanism that has not yet been evaluated in the context of Nigeria.

Here, we leveraged the baseline (2014) and endline (2017) facility surveys, which included documentation of product availability as well as supply chain management practices and availability of relevant infrastructure. (33) We quantified the effect of DFF and PBF on product availability above and beyond underlying secular trends, examined whether there were differences in supply chain performance between DFF and PBF facilities, and studied which clinical areas benefited most in order to better understand what other barriers exist that could benefit from further study.

While study data is from 2014-2017, the Nigerian government adopted the decentralized financing approach in the implementation of the Basic Health Care Provision Fund (BHCPF), which has been scaled across the 36 states and the Federal Capital Territory since then. (34) Findings from this study can provide useful information to support the ongoing operational implementation choices of the BHCPF and can be helpful to other countries experiencing similar supply chain challenges.

## Materials & Methods

### Data

We conduct secondary data analysis on the NSHIP facility survey data, which was collected by independent observers who visited every facility in 2014 (“baseline”) and again in 2017 (“endline) as part of the project evaluation. We leverage the facility survey data, which includes detailed reporting on availability of individual products at the time of the survey as well as questions on supply chain practices and cold chain equipment.

All data has limitations and in this case, the primary assumption that we make is that “NA” values are considered zeroes (i.e., not in stock). This skip pattern in the survey occurred if a facility did not deliver a particular service. For example, if they stated that they do not provide malaria care, then their stock levels for specific malaria treatment drugs were presumably zero, but this was not directly recorded. We believe this to be a reasonable approximation of reality and limits our loss of sample size or risk of a biased sample.

### Outcome measures

For the purposes of this paper, we consider two primary outcome measures.

First is the total number of essential drugs available on the day of the survey, by facility. We look at this in aggregate and then also examine subsets relevant for services including routine immunization (RI) and obstetrics. Where it provides useful insights, we report how individual products changed between baseline and endline.

Where possible, we report outcomes by clinical service area to ascertain whether certain product categories responded more to the intervention than others.

To better understand how the mechanisms by which DFF and PBF under the NSHIP interventions may have affected product availability, we examined corresponding changes in supply chain functionality. These included cold chain items (main unit thermometers, cold storage units, cold boxes, and vaccine carriers) and management practices (bringing in money from drug sales, and logging refrigerator temperatures). We also examined immunization-related indicators of staff activities (number of outreach sessions in the last month, and number of trips to the LGA in the last month).

### Analysis approach

We assessed the effectiveness of DFF and PBF by comparing baseline and endline performance for control versus intervention groups. The original study’s design was to assess whether performance-based financing (PBF) outperformed DFF, and how both compared to a continuation of business as usual (control), so we further break down the results control vs. DFF vs. PBF in the endline comparisons to reflect the reality that these subsets of intervention facilities had different interventions and funding levels available to them.

We report descriptive statistics for our two primary outcome measures and results from Wilcox tests on the difference in means or Mood’s test for difference in medians as appropriate, looking at each set of comparison groups. These include:

- Control facilities – baseline vs. endline,
- DFF and PBF facilities – baseline vs. endline,
- Endline – control vs. intervention, and
- Endline – DFF vs. PBF.

To examine the effects on supply chain enablers and functionality, we did three things. First, we reported average values for each state and note the directional change across each of these measures. Second, we reported statistical significance for tests comparing baseline to endline. Third, we ran a difference-in-difference (DiD) linear mixed effects regression (LMER) where product availability was the outcome, enablers and functionality were predictors, study arm was treated as a fixed effect, state is a random effect, and we included the interaction between study arm and time.

## Results

### Changes in product availability

We see that product availability for essential medicines and all products were similar at baseline but increased much more in intervention facilities, significantly outperforming control at endline. PBF did modestly better than DFF. (Figure 1) We note that this approach to summarizing availability treats this as a binary measure (available or not), without regard for quantity on the shelf exceeding one.

**Figure 1.**
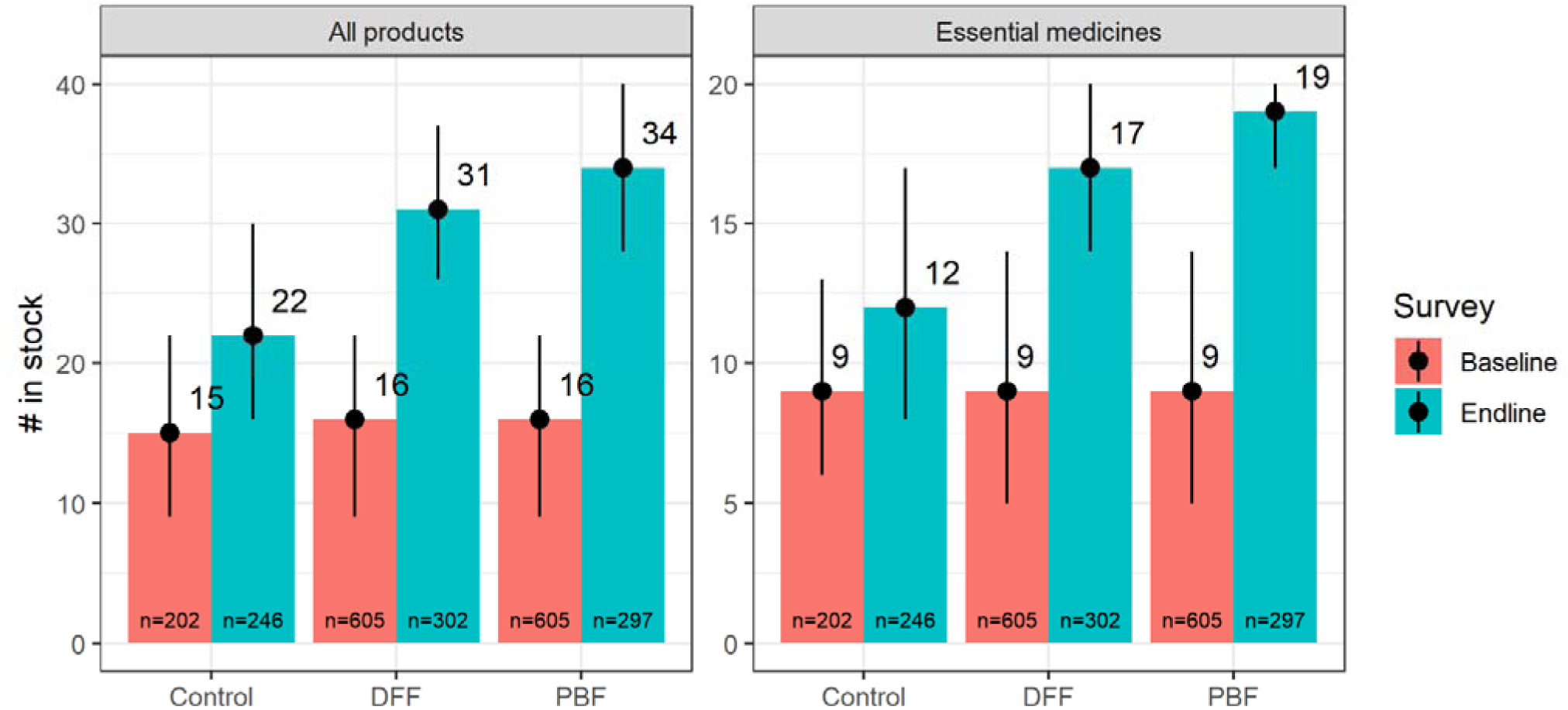
Descriptive statistics for the two primary outcome measures. Bar heights are the median, with the 25^th^ and 75^th^ percentiles shown as uncertainty lines. Essential medicines include all items listed by the WHO that were included in the facility survey, so the maximum possible score is 29. Control includes facilities in selected LGAs in Benue, Ogun, and Taraba. Intervention includes all PBF and DFF facilities in Adamawa, Ondo, and Nasarawa. DFF = direct facility financing. PBF = performance-based financing. Baseline = 2014. Endline = 2017. n=sample size.

Looking at availability by product category, we split out three subsets of products to examine them in more detail: routine immunization (RI), family planning (FP), and obstetrics. When we do so, we note that all facility groups (control, DFF and PBF) experienced increases in total products on shelf. However, these gains were more significant overall for intervention facilities (DFF +12 and PBF +18) than control (+3). Looking at specific product families, these increases were not driven by services that are managed in a vertical way, such as routine immunization or family planning. Instead, the additional gains in DFF and PBF facilities came from products like antibiotics, diagnostics, and other drug families. (Table 1)

**Table 1.** Results from Wilcox tests for difference in means, total number of products in stock today. Values reported are the average (mean) values and the p-value is the result from the Wilcox test. RI = routine immunization. FP = family planning. DFF = facilities assigned to direct facility financing. PBF = facilities assigned to performance-based financing. Intervention category includes both DFF and PBF facilities. Baseline = 2014 facility survey. Endline = 2017 facility survey. Difference values are shaded if they were significant, according to the size of the delta.

| Data set | Comparison | Product (max #) | Value 1 (mean) | Value 2 (mean) | Difference | p-value |
| --- | --- | --- | --- | --- | --- | --- |
| Control | Baseline (value 1) vs<br>Endline (value 2) | All products (66) | 14.5 | 17.2 | 2.7 | < 0.001 |
|  |  | RI (9) | 1.1 | 4.4 | 3.3 | < 0.001 |
|  |  | FP (5) | 1.8 | 3.3 | 1.5 | < 0.001 |
|  |  | Obstetric (8) | 2.4 | 2.9 | 0.5 | < 0.001 |
|  |  | Antibiotics (13) | 5.0 | 5.0 | 0 | 0.90 |
| DFF | Baseline (value 1) vs<br>Endline (value 2) | All products (66) | 13.7 | 26.0 | 12.3 | < 0.001 |
|  |  | RI (9) | 2.4 | 4.5 | 2.1 | < 0.001 |
|  |  | FP (5) | 2.0 | 3.2 | 1.2 | < 0.001 |
|  |  | Obstetric (8) | 2.6 | 4.0 | 1.4 | < 0.001 |
|  |  | Antibiotics (13) | 4.5 | 7.2 | 2.7 | <0.001 |
| PBF | Baseline (value 1) vs<br>Endline (value 2) | All products (66) | 13.7 | 31.6 | 17.9 | < 0.001 |
|  |  | RI (9) | 2.4 | 4.7 | 2.3 | < 0.001 |
|  |  | FP (5) | 2.0 | 3.5 | 1.5 | < 0.001 |
|  |  | Obstetric (8) | 2.6 | 4.1 | 1.5 | < 0.001 |
|  |  | Antibiotics (13) | 4.5 | 8.7 | 4.2 | <0.001 |
| Endline | Control (value 1) vs<br>DFF+PBF (value 2) | All products (66) | 17.2 | 29.2 | 12.0 | < 0.001 |
|  |  | RI (9) | 4.4 | 4.6 | 0.2 | 0.70 |
|  |  | FP (5) | 3.3 | 3.4 | 0.1 | 0.40 |
|  |  | Obstetric (8) | 2.9 | 4.0 | 1.1 | < 0.001 |
|  |  | Antibiotics (13) | 5.0 | 8.1 | 3.1 | <0.001 |
| Endline | DFF (value 1) vs<br>PBF (value 2) | All products (66) | 26.0 | 31.6 | 5.6 | < 0.001 |
|  |  | RI (9) | 4.5 | 4.7 | 0.2 | 0.30 |
|  |  | FP (5) | 3.2 | 3.5 | 0.3 | < 0.001 |
|  |  | Obstetric (8) | 4.0 | 4.1 | 0.1 | 0.60 |
|  |  | Antibiotics (13) | 7.2 | 8.7 | 1.5 | <0.001 |

We ran a DiD regression on total products and total essential products and found that the intervention effect of DFF and PBF had a significant and positive effect on product availability on the day of the endline survey. At baseline, facilities in the study arm had 2.7 more products in stock on the day of the survey than control facilities, and the same number of essential medicines. The average change in the control arm, and the assumed overall time trend, was 0 in both cases. The interaction term of study arm and time, the key model parameter which captures the effect of the intervention over the course of the study, was significant for both the total number of products in stock and the number of essential medicines in stock. These effect sizes were +12.8 and +7.8 products on shelf, respectively, and estimate the increase relative to control. (Table 2) The maximum number of products being tracked was 66 and 29, so these effect sizes were equivalent to a 19.4% and 26.9% increase in the number of products available, respectively.

**Table 2.** Regression results estimating the effect of PBF and DFF on product availability. Effect size is the number of additional products on the shelf resulting from a particular variable. In this table, the interaction term (study arm*time) is the key variable and its significance indicates the impact of the study on product availability. The total possible number of products in stock was 66. The number of potential essential medicines in stock was 29. Cells are shaded according to p-value.

| Variable | Total # products in stock |  | # essential medicines in stock |  |
| --- | --- | --- | --- | --- |
|  | Effect size | p-value | Effect size | p-value |
| Study arm<br>(intervention vs. control) | 2.7 | 0.004 | 0.4 | 0.503 |
| Time<br>(endline vs baseline) | -0.8 | 0.309 | -0.6 | 0.157 |
| Study arm * Time | 12.8 | <0.001 | 7.8 | <0.001 |
| Sample Size | 1752 |  | 1752 |  |

### Supply chain enablers and management practices

On average, both control and intervention facilities had improvements in supply chain enablers and functionality, but the gains were larger in the intervention arm. On average, PBF facilities had somewhat better performance than DFF. Most notably, the average control facility still had no refrigerator by endline, but the average intervention facility did have one, allowing them to safely store vaccines on site. There were also larger increases in ice pack availability and the frequency of temperature checks. Staff in intervention facilities increased their outreach services and frequency of their trips to the LGA to pick up supplies more than control facilities, possibly reflecting how increased money was spent on logistics. Lastly, intervention facilities saw a more dramatic increase in drug sales for cost recovery and additional revenues for the facilities. (Figure 2)

**Figure 2.**
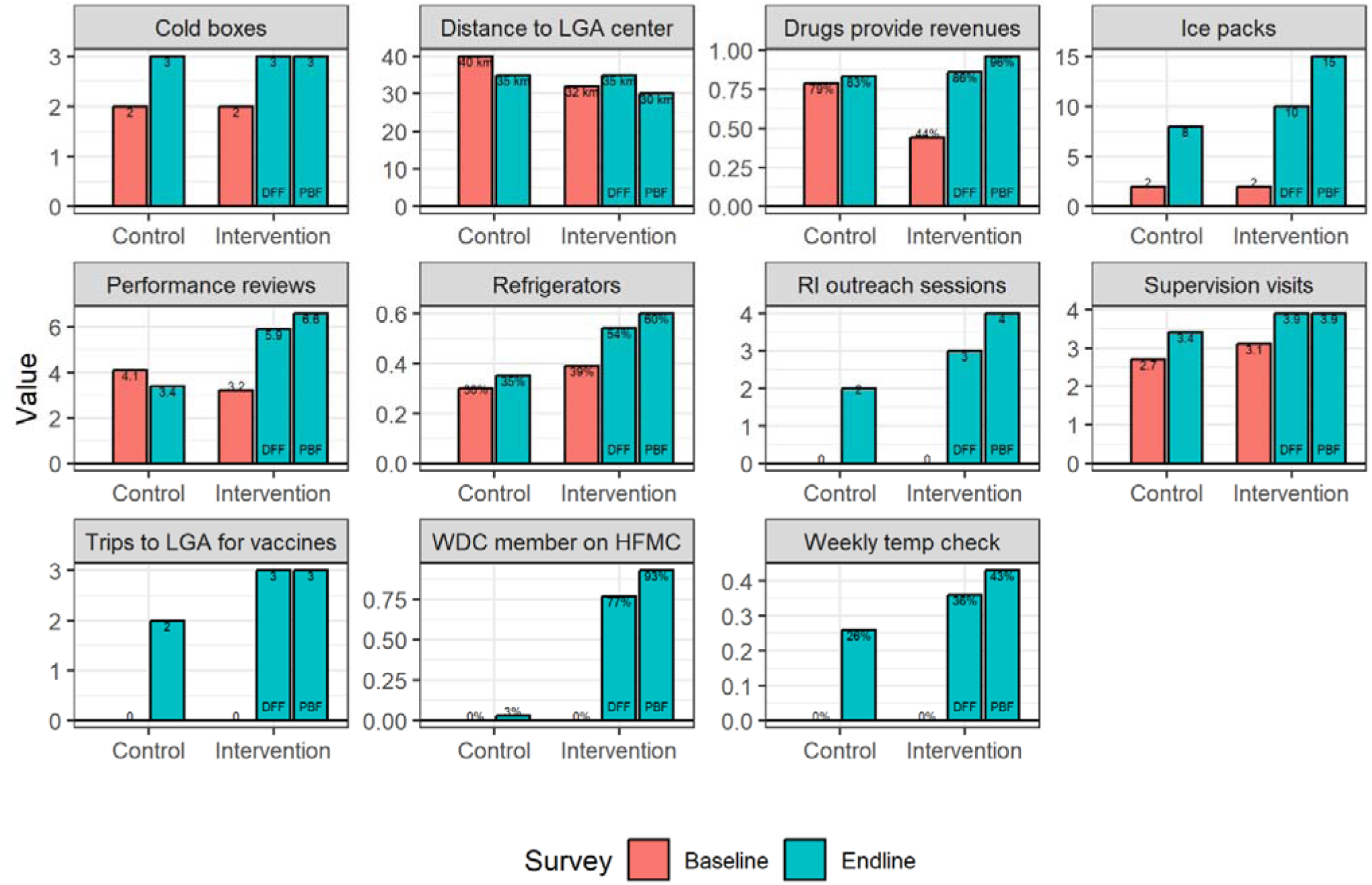
Change in supply chain enablers and functionality between baseline (2014) and endline (2017). Data was collected by independent observers as part of facility surveys. Some measures are self-reported by the facility based on their documentation, such as outreach sessions and trips to the LGA. RI = routine immunization. DFF = direct facility financing. PBF = performance-based financing. LGA = local government administration. Intervention values are shaded if they were significantly different than the control arm from the same survey.

Statistically speaking (using Mood’s test for differences in median values), at endline, intervention facilities had higher performance than control facilities (p<0.01) for all of the following: ice pack availability, refrigerators, frequency of temperature checks, number of outreach sessions, number of trips to LGA, drug sales providing revenues, frequency of performance review, and having a ward development committee (WDC) member on the health facility management committee (HFMC). While PBF median values were always the same or better than DFF, the only measures for which the difference was statistically significant were ‘drug sales providing revenues to the facility’ and ‘having a WDC member on the HFMC’ (p<0.001).

The percentage of facilities with one or more functional refrigerators increased by only 5% (non-significant) in the control arm, while DFF and PBF facilities increased by +15% and 21%, respectively. This was a statistically significant difference and represents an important programmatic difference in terms of supply chain capacity.

Further, DiD LMER results found that many of these enablers and functionality measures were associated with an increase in product availability in the facility. The model results indicate that a facility in the intervention arm of the study had a net increase of 5.6 essential medicines and 8.2 total products between baseline and endline. All but one of the enablers were statistically significant in predicting the number of available products in stock on the day of the survey. (Table 3) Of note, routine cold chain maintenance (as measured by temperature checks), conducting outreach sessions, using drug sales as a facility revenue source (presumably to replace drugs that are consumed), frequent supervision and evaluation were all positive. Travel time to the LGA distribution center was negatively associated with performance – i.e., facilities that had to go farther to replenish products had more stock-outs.

**Table 3.**
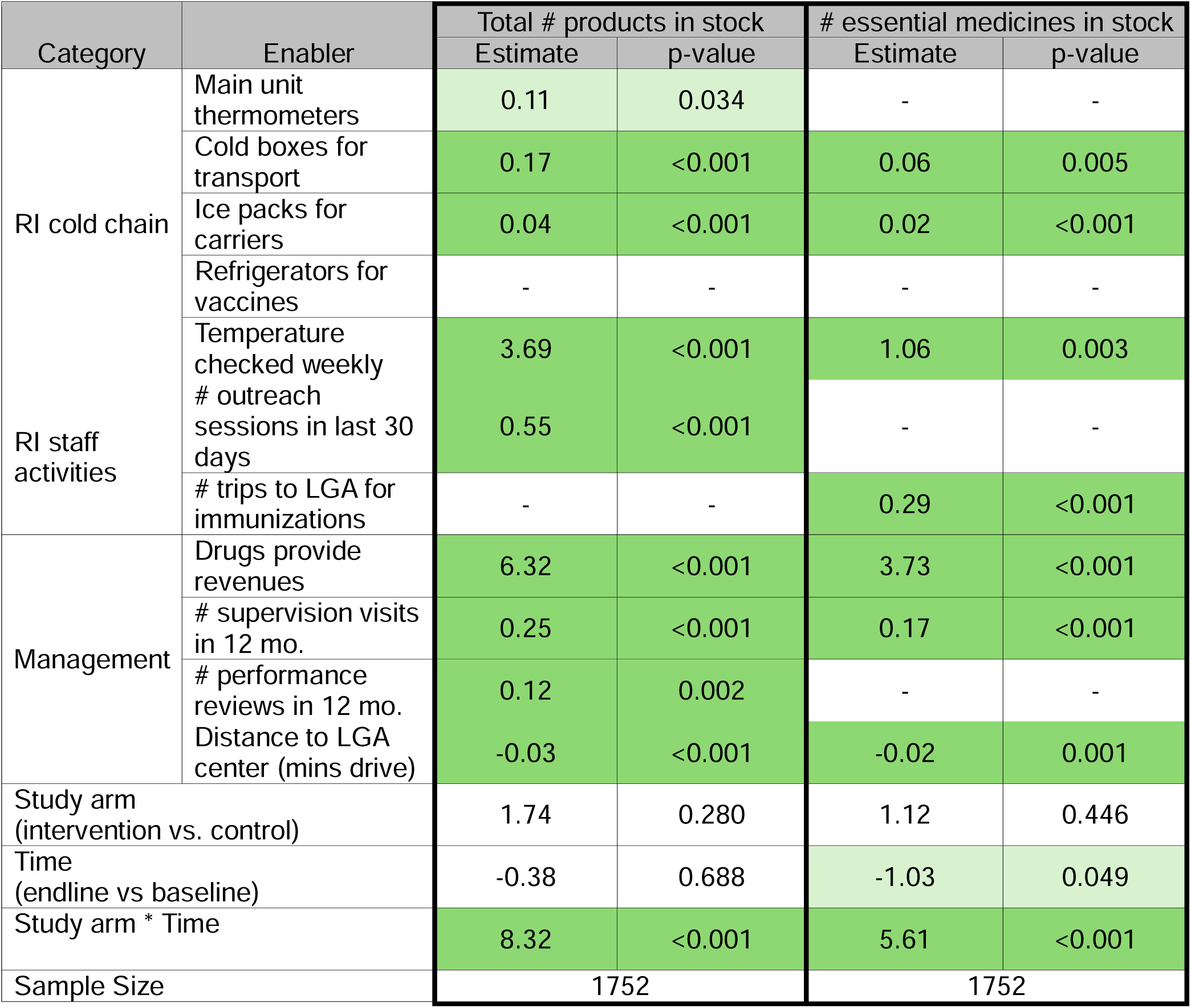
Difference-in-difference regression results. Two outcome measures were tested: the total number of products in stock and the number of essential medicines in stock; both were tested as a robustness check on the results. Reported values are the coefficient estimate and the calculated p-value. Shading is to make the table easier to read, with parameters that were significant at p < 0.05 in light green and p < 0.001 in dark green.

There were additional variables of interest pertaining to supply chain management capacity, but due to data limitations, a DiD regression could not be performed. However, an LMER model using only the endline data showed that several additional variables were statistically and positively associated with product availability, including: having a separate room for the pharmacy, having a pharmacy that has lockable doors and windows for security, maintaining stock cards or a stock register showing the security stock level, all drugs and medical consumables being NAFDAC certified, and having a member of the WDC on the HFMC. Having a trained pharmacist on staff was positively associated with increased product availability, but not significant (coefficient 1.83, p-value 0.21). (See Supplementary Table S2).

## Discussion

### Large Overall Improvements

DFF and PBF in Nigeria resulted in an increase in availability of products and drugs, both in the DFF and PBF study arms. While the control facilities also saw some modest increases during the same timeframe, this was mostly in vertically managed programs such as RI. The DFF and PBF facilities had significantly larger gains across the product portfolio. The modeled estimate of impact of DFF and PBF was +12.8 additional products, of which +7.8 were essential medicines. In absolute terms, the increase in total product availability for DFF was +12 and PBF +18, compared to control +3, representing a 17% and 27% (percentage point) reduction in stock-outs. This is better than the 7% improvement in Tanzania after implementing a logistics management system and in line with the results from a fundamental redesign of Zambia’s supply chain, but with less intensive intervention or disruption.

### Mechanisms

To better understand the mechanism by which this improvement occurred, we examined several enablers and functionality of the supply chain. We find that again, the PBF and DFF districts outperformed control in terms of improvements in the state of the enablers, e.g., with more frequent trips to pick up supplies, and weekly refrigerator temperature checks. Of note is that the median for refrigerator availability in intervention facilities increased from zero to one, a programmatically important step-change in equipment availability.

At the same time, the number of vaccination outreach sessions increased as well, suggesting that these changes had a meaningful impact on facility operations and service availability, likely driving the previously described increase in immunization coverage. (32) The reported reduction in travel time to the LGA store and increased frequency of those trips both would be expected to increase product availability, suggesting that facilities in part used their additional funds to cover transportation costs (e.g., fuel) in order to reduce stock-outs.

Difference-in-difference regression analysis showed that these enablers were significantly associated with higher product availability (both total and essential products only).

Of particular interest was the strong relationship between facilities using drug sales as a revenue source, which had the largest effect size, aside from study arm itself. This suggests that the total budget available to facilities is an important driver of increased product availability and ensuring adequate funding would likely have long-term benefits.

The next largest effects came from monthly supervision visits (coefficient of 0.25*12 = +3.0) and routine temperature checks (+ 3.7). These are both equivalent to more than a third of the effect of being in the study, suggesting that good supply management practices at the facility level are important and should be prioritized for investment.

Data on additional enablers related to facility infrastructure were available only in the endline survey, so while we could not do a longitudinal regression, we instead conducted a regression analysis only on intervention facilities, to see which components may contribute to increased product and drug availability.

In doing so, we found that having a well-equipped facility (e.g., secure storage), NAFDAC-certified products, and having a WDC on the HFMC had substantial positive effects on product availability. This is important to note since the requirement for certification would in fact limit the pool of potential suppliers, so having such a large positive effect (+6.8) would imply that NAFDAC requirements do not hinder availability. Having a separate and secure pharmacy also had a large (+7.3) effect, suggesting that safe storage can reduce pilferage and potentially increase facility staff confidence in purchasing stock that will be available for users when it is needed. Additionally, the size of the effect of a WDC on the management committee was sizeable at + 4.8 total products, emphasizing the importance of good community engagement in long-term management practices.

These effects are large compared to the coefficient from being in the PBF arm, suggesting that within intervention facilities at endline, the things that mattered most were specific practices and policies, and facility infrastructure (presumably purchased through NSHIP incremental funds), not only the fact of being in the study itself.

From this, we conclude that DFF and PBF had a meaningful and important impact on supply chain equipment, drug availability, and operational choices that drove the previously observed increases in service coverage.

This is an important conclusion in itself, but we also note that DFF and PBF were intended to improve overall facility performance, not aiming for supply chain specifically, so the large effect on product availability was not a guarantee. However, given the results, we conclude that by giving facilities modest funding and the autonomy to make decisions, they were able to effectively address their supply chain issues in ways that worked for them, and ultimately increased product availability. This was a pull system, not push, a fundamentally different way of working than business as usual had been, a finding that raises the question of whether central medical stores are necessary. It also implies that bringing in the private sector, when properly certified for quality products, may be an effective supply chain component.

### Price & Quality

Two major objections are typically raised to using DFF or PBF to improve supply chains: the price of products and the quality of the medicines.

The evaluation of NSHIP did not collect data directly on prices paid for essential drugs, but an analysis carried out at the same time found that median price difference between government stores and private suppliers was 79%, i.e., government procured medicines were considerably more expensive than those in the private sector. (35)

On the issue of quality, we find that NAFDAC approved medicines were more likely in stock at DFF and PBF facilities (compared to control) and that pharmacy management was considerably higher (see supplemental results). There have been concerns expressed about the prevalence of counterfeit drugs in Nigeria (12) but a joint NAFDAC-United States Pharmacopeia study in 2017 looking at malaria medicines sampled from around the country found only 0.9% had no active ingredient and another 3.4% of the sample were substandard (failed dissolution or assay tests). (14)

### Expansion of DFF & PBF

Many low and middle income (LMIC) countries including Nigeria have either adopted or scaled up direct facility financing (plus performance-based financing) as a health financing strategy. (36,37) Correspondingly, multiple innovations are being implemented to adjust or strengthen the health system especially in areas of governance and leadership where there has been greater involvement of the communities in the management of health facilities and in extension of public financial management system to the facility level. (38) This paper makes the case for rethinking the old, centralized logistic and supply chain (LSC) management (push system) for greater efficiency to ensure availability of medicines and consumables at the last mile. In NSHIP, there was no investment to central medical stores, there was shared distribution cost, and crowding in of the private sector at the local level in sort of public private partnership (PPP). To further improve LSC system, Tanzania introduced prime vendors into its health system (39), just as in the case of NSHIP in Nigeria and Liberia uses a contractual framework agreement (CFA) ‘between the counties/MOH and the pharmaceutical wholesaler defines fixed prices for a list of essential medicines, inclusive of delivery to the county depot within 10 days of receiving the purchase order and stipulates that all medicines should be registered by the Liberia Medicines and Health Products Regulatory Authority’. (40)

The results presented here call for different roles for the Ministry of Health in the procurement, storage and distribution of medicines and consumables to address the inherent challenges and to align with decentralization policies of governments of LMICs including Nigeria; with the goal of deepening public-private partnerships (PPPs) in supply chains to improve service delivery. However, selection of reputable and competent vendors, and sticking to the contractual agreement is critically important.

## Conclusion

We find that the DFF and PBF interventions, which consisted primarily of incremental funding and facility autonomy to spend it as needed, had a positive effect on the in-stock availability of medicines and products. While the study was not designed specifically to target supply chain performance, it nevertheless had a meaningful impact, suggesting that putting money into the hands of local facilities is an effective intervention. During the study, the money seems to have been spent on improving infrastructure (e.g., secure rooms & refrigerators), addressing operational needs to improve re-supply, and purchasing NAFDAC-certified drugs. The effectiveness of the intervention also raises questions about the need for central stores and whether there is a larger role for private distributors to play. As a result of these findings, decision makers should consider facility-level financing as a potential intervention, alongside policy reforms that would open up supply chains and improve product availability.

## Data Availability

No new data was produced during this study. All data used for the analysis are available from the World Bank's microdata library upon request.

https://microdata.worldbank.org/index.php/catalog/4042

## Acknowledgements

The authors would like to thank Eeshani Kandpal for supporting the data cleaning and access to the full datasets that made this work possible.

