## Supplement 1 for "How decentralized facility financing improved supply chains and product availability in primary healthcare centers, a randomized trial in Nigeria"

### **Supplementary Materials**

Table S1. Change in supply chain enablers and functionality. Arrows indicate values at different time points such as: baseline 🡪 endline. Cells are colored such that light green indicates an increase, dark green indicates an increase where the starting value was zero, and light orange indicates a decrease.

|  |  | Control | | | Intervention | | |
| --- | --- | --- | --- | --- | --- | --- | --- |
|  | Enabler | Benue | Ogun | Taraba | Adamawa | Nasarawa | Ondo |
| Cold chain | Main unit thermometers | Median 0🡪0 | Median 0🡪0 | Median 0🡪0 | Median 0🡪0 | Median 0🡪0 | Median 0🡪0 |
|  | Cold boxes for transport | Median 3🡪5 | Median 2🡪3 | Median 2🡪2 | Median 2🡪3 | Median 2🡪2 | Median 4🡪4 |
|  | Ice packs for carriers | Median 4🡪10 | Median 1🡪8 | Median 0🡪7 | Median 0🡪10 | Median 2🡪6 | Median 10🡪24 |
|  | Refrigerators for vaccines | Median 0🡪0 | Median 0🡪0 | Median 0🡪0 | Median 0🡪1 | Median 0🡪1 | Median 0🡪1 |
|  | Temperature checked weekly | 0% 🡪 11% | 0% 🡪 28% | 0% 🡪 36% | 0% 🡪 49% | 0% 🡪 34% | 0% 🡪 30% |
| Staff activities | 1+ outreach in last 30 days | 0% 🡪 86% | 0% 🡪 94% | 0% 🡪 92% | 0% 🡪 92% | 0% 🡪 87% | 0% 🡪 97% |
|  | # trips to LGA for supplies | Median  0 🡪 1 | Median  0 🡪 2 | Median  0 🡪 3 | Median  0 🡪 4 | Median  0 🡪 2 | Median  0 🡪 2 |
| Financing | Drugs provide revenues | 87% 🡪 90% | 70% 🡪 89% | 83% 🡪 71% | 53% 🡪 92% | 82% 🡪 98% | 11% 🡪 88% |
|  | Insurance $ used to purchase drugs | 0% 🡪 0% | 0% 🡪 2% | 0% 🡪 1% | 0% 🡪 0% | 0% 🡪 2% | 21% 🡪 6% |
|  | Sample size | 63/73 | 80/81 | 59/92 | 266/338 | 128/131 | 211/230 |

Table S2. Endline-only regression results, intervention facilities only. Two outcome measures were tested: the total number of products in stock and the number of essential medicines in stock; both were tested as a robustness check on the results. All management capacity values were binary (yes/no). Results values reported in the table are the coefficient estimate and the calculated p-value. Shading is to make the table easier to read, with parameters that were significant at p < 0.05 in light green and p < 0.001 in dark green.

| Category | Enabler | Total # products in stock | | # essential medicines in stock | |
| --- | --- | --- | --- | --- | --- |
|  |  | Estimate | p-value | Estimate | p-value |
| RI cold chain | Main unit thermometers | 0.93 | 0.011 | - | - |
|  | Cold boxes for transport | 0.18 | 0.005 | 0.07 | 0.024 |
|  | Ice packs for carriers | 0.02 | 0.012 | 0.01 | 0.013 |
|  | Refrigerators for vaccines | 0.86 | <0.001 | 0.37 | 0.004 |
|  | Temperature checked weekly | 1.90 | 0.004 | - | - |
| RI staff activities | # outreach sessions in last 30 days | 0.42 | 0.007 | - | - |
|  | # trips to LGA for immunizations | - | - | 0.25 | <0.001 |
| Financing | Drugs provide revenues | 5.62 | <0.001 | 3.41 | <0.001 |
| Pharmacy capacity | Trained pharmacist on staff | - | - | - | - |
|  | Separate pharmacy storage room | 3.49 | 0.001 | 1.94 | <0.001 |
|  | Pharmacy room is securable | 3.78 | 0.003 | 1.96 | <0.001 |
|  | Maintain stock cards or register | 3.58 | <0.001 | 2.08 | 0.003 |
|  | All consumables NAFDAC certified | 6.80 | <0.001 | 4.71 | <0.001 |
|  | Purchase from certified distributor | - | - | - | - |
|  | All drugs prescribed in generic form | - | - | - | - |
| Management capacity | # supervision visits in 12 mo. | 0.22 | 0.010 | - | - |
|  | # performance reviews in 12 mo. | - | - | - | - |
|  | Distance to LGA center (mins drive) | - | - | - | - |
|  | WDC member on the HFMC | 4.79 | <0.001 | 2.69 | <0.001 |
| Study arm  (PBF vs DFF) | | 1.95 | 0.002 | 1.57 | <0.001 |

Table S3. Wilcoxin tests for all product groups within the full product portfolio.

| Data set | Comparison | Product (max #) | Value 1 (mean) | Value 2 (mean) | Difference | p-value |
| --- | --- | --- | --- | --- | --- | --- |
| Control | Baseline vs Endline | All products (66) | 14.5 | 17.2 | 2.7 | < 0.001 |
|  |  | RI (9) | 1.1 | 4.4 | 3.3 | < 0.001 |
|  |  | FP (5) | 1.8 | 3.3 | 1.5 | < 0.001 |
|  |  | Obstetric (8) | 2.4 | 2.9 | 0.5 | < 0.001 |
|  |  | Antibiotics (13) | 5.0 | 5.0 | - | 0.90 |
|  |  | Cardiovascular (4) | 0.3 | 1.1 | 0.8 | < 0.001 |
|  |  | Diagnostics (5) | 1.5 | 2.2 | 0.7 | < 0.001 |
|  |  | General (4) | 1.4 | 1.8 | 0.4 | < 0.001 |
|  |  | Malaria (5) | 1.1 | 1.8 | 0.7 | 0.1 |
|  |  | Specific (9) | 1.8 | 2.5 | 0.7 | < 0.001 |
|  |  | TB (7) | 0.8 | 2.2 | 1.4 | < 0.001 |
| DFF | Baseline vs Endline | All products (66) | 13.7 | 26.0 | 12.3 | < 0.001 |
|  |  | RI (9) | 2.4 | 4.5 | 2.1 | < 0.001 |
|  |  | FP (5) | 2.0 | 3.2 | 1.2 | < 0.001 |
|  |  | Obstetric (8) | 2.6 | 4.0 | 1.4 | < 0.001 |
|  |  | Antibiotics (13) | 4.5 | 7.2 | 2.7 | <0.001 |
|  |  | Cardiovascular (4) | 0.3 | 1.2 | 0.9 | < 0.001 |
|  |  | Diagnostics (5) | 1.8 | 3.2 | 1.4 | < 0.001 |
|  |  | General (4) | 1.6 | 2.4 | 0.8 | < 0.001 |
|  |  | Malaria (5) | 1.4 | 1.8 | 0.4 | < 0.001 |
|  |  | Specific (9) | 1.8 | 3.2 | 1.4 | < 0.001 |
|  |  | TB (7) | 0.9 | 2.9 | 2.0 | < 0.001 |
| PBF | Baseline vs Endline | All products (66) | 13.7 | 31.6 | 17.9 | < 0.001 |
|  |  | RI (9) | 2.4 | 4.7 | 2.3 | < 0.001 |
|  |  | FP (5) | 2.0 | 3.5 | 1.5 | < 0.001 |
|  |  | Obstetric (8) | 2.6 | 4.1 | 1.5 | < 0.001 |
|  |  | Antibiotics (13) | 4.5 | 8.7 | 4.2 | <0.001 |
|  |  | Cardiovascular (4) | 0.3 | 1.2 | 0.9 | < 0.001 |
|  |  | Diagnostics (5) | 1.8 | 3.7 | 2.9 | < 0.001 |
|  |  | General (4) | 1.6 | 2.9 | 1.3 | < 0.001 |
|  |  | Malaria (5) | 1.4 | 1.9 | 0.5 | < 0.001 |
|  |  | Specific (9) | 1.8 | 3.6 | 1.8 | < 0.001 |
|  |  | TB (7) | 0.9 | 3.3 | 2.4 | < 0.001 |
| Endline | Control vs Intervention | All products (66) | 17.2 | 29.2 | 12.0 | < 0.001 |
|  |  | RI (9) | 4.4 | 4.6 | 0.2 | 0.70 |
|  |  | FP (5) | 3.3 | 3.4 | 0.1 | 0.40 |
|  |  | Obstetric (8) | 2.9 | 4.0 | 1.1 | < 0.001 |
|  |  | Antibiotics (13) | 5.0 | 8.1 | 3.1 | <0.001 |
|  |  | Cardiovascular (4) | 1.1 | 1.2 | 0.1 | 0.6 |
|  |  | Diagnostics (5) | 2.2 | 3.5 | 1.3 | < 0.001 |
|  |  | General (4) | 1.8 | 2.7 | 0.9 | < 0.001 |
|  |  | Malaria (5) | 1.8 | 1.9 | 0.1 | 0.02 |
|  |  | Specific (9) | 2.5 | 3.4 | 0.9 | < 0.001 |
|  |  | TB (7) | 2.2 | 3.1 | 0.9 | < 0.001 |
| Endline | DFF vs PBF | All products (66) | 26.0 | 31.6 | 5.6 | < 0.001 |
|  |  | RI (9) | 4.5 | 4.7 | 0.2 | 0.30 |
|  |  | FP (5) | 3.2 | 3.5 | 0.3 | < 0.001 |
|  |  | Obstetric (8) | 4.0 | 4.1 | 0.1 | 0.60 |
|  |  | Antibiotics (13) | 7.2 | 8.7 | 1.5 | <0.001 |
|  |  | Cardiovascular (4) | 1.2 | 1.2 | - | 0.3 |
|  |  | Diagnostics (5) | 3.2 | 3.7 | 0.5 | < 0.001 |
|  |  | General (4) | 2.4 | 2.9 | 0.5 | < 0.001 |
|  |  | Malaria (5) | 1.8 | 1.9 | 0.1 | 0.2 |
|  |  | Specific (9) | 3.2 | 3.6 | 0.4 | 0.005 |
|  |  | TB (7) | 2.9 | 3.3 | 0.4 | 0.1 |

Table S4. Wilcoxin tests for all available product groups within the essential medicines product portfolio.

| Data set | Comparison | Product (max #) | Value 1 (mean) | Value 2 (mean) | Difference | p-value |
| --- | --- | --- | --- | --- | --- | --- |
| Control | Baseline vs Endline | All essential (29) | 8.8 | 9.1 | 0.3 | 0.6 |
|  |  | RI (0) | - | - | - | - |
|  |  | FP (5) | 1.8 | 3.3 | 1.5 | < 0.001 |
|  |  | Obstetric (2) | 0.9 | 1.2 | 0.3 | < 0.001 |
|  |  | Antibiotics (10) | 4.6 | 4.6 | - | 0.9 |
|  |  | Cardiovascular (3) | 0.3 | 1.1 | 0.8 | < 0.001 |
|  |  | Diagnostics (0) | - | - | - | - |
|  |  | General (4) | 1.4 | 1.8 | 0.4 | < 0.001 |
|  |  | Malaria (2) | 0.2 | 0.2 | - | 0.1 |
|  |  | Specific (3) | 0.7 | 0.9 | 0.2 | 0.002 |
|  |  | TB (0) | - | - | - | - |
| DFF | Baseline vs Endline | All essential (29) | 8.2 | 14.3 | 6.1 | < 0.001 |
|  |  | RI (0) | - | - | - | - |
|  |  | FP (5) | 2.0 | 3.2 | 1.2 | < 0.001 |
|  |  | Obstetric (2) | 1.1 | 1.5 | 0.4 | < 0.001 |
|  |  | Antibiotics (10) | 4.1 | 6.4 | 2.3 | < 0.001 |
|  |  | Cardiovascular (3) | 0.3 | 1.2 | 0.9 | < 0.001 |
|  |  | Diagnostics (0) | - | - | - | - |
|  |  | General (4) | 1.6 | 2.4 | 0.8 | < 0.001 |
|  |  | Malaria (2) | 0.2 | 0.2 | - | 0.7 |
|  |  | Specific (3) | 0.8 | 1.4 | 0.6 | < 0.001 |
|  |  | TB (0) | - | - | - | - |
| PBF | Baseline vs Endline | All essential (29) | 8.2 | 17.8 | 9.6 | < 0.001 |
|  |  | RI (0) | - | - | - | - |
|  |  | FP (5) | 2.0 | 3.5 | 1.5 | < 0.001 |
|  |  | Obstetric (2) | 1.1 | 1.7 | 0.6 | < 0.001 |
|  |  | Antibiotics (10) | 4.1 | 7.7 | 3.6 | < 0.001 |
|  |  | Cardiovascular (3) | 0.3 | 1.1 | 0.8 | < 0.001 |
|  |  | Diagnostics (0) | - | - | - | - |
|  |  | General (4) | 1.6 | 2.9 | 1.3 | < 0.001 |
|  |  | Malaria (2) | 0.2 | 0.2 | - | 0.02 |
|  |  | Specific (3) | 0.8 | 1.6 | 0.8 | < 0.001 |
|  |  | TB (0) | - | - | - | - |
| Endline | Control vs Intervention | All essential (29) | 9.1 | 16.3 | 7.2 | < 0.001 |
|  |  | RI (0) | - | - | - | - |
|  |  | FP (5) | 3.3 | 3.4 | 0.1 | 0.3 |
|  |  | Obstetric (2) | 1.2 | 1.6 | 0.4 | < 0.001 |
|  |  | Antibiotics (10) | 4.6 | 7.1 | 2.5 | < 0.001 |
|  |  | Cardiovascular (3) | 1.1 | 1.1 | - | 0.4 |
|  |  | Diagnostics (0) | - | - | - | - |
|  |  | General (4) | 1.8 | 2.7 | 0.9 | < 0.001 |
|  |  | Malaria (2) | 0.2 | 0.2 | - | 0.8 |
|  |  | Specific (3) | 0.9 | 1.5 | 0.6 | < 0.001 |
|  |  | TB (0) | - | - | - | - |
| Endline | DFF vs PBF | All essential (29) | 14.3 | 17.8 | 3.5 | < 0.001 |
|  |  | RI (0) | - | - | - | - |
|  |  | FP (5) | 3.2 | 3.5 | 0.3 | < 0.001 |
|  |  | Obstetric (2) | 1.5 | 1.7 | 0.2 | 0.008 |
|  |  | Antibiotics (10) | 6.4 | 7.7 | 1.3 | < 0.001 |
|  |  | Cardiovascular (3) | 1.2 | 1.1 | -0.1 | 0.3 |
|  |  | Diagnostics (0) | - | - | - | - |
|  |  | General (4) | 2.4 | 2.9 | 0.5 | < 0.001 |
|  |  | Malaria (2) | 0.2 | 0.2 | - | 0.1 |
|  |  | Specific (3) | 1.4 | 1.6 | 0.2 | 0.01 |
|  |  | TB (0) | - | - | - | - |
